# Predictive models for hospitalization and mortality in dengue using SINAN data: study protocol for development, temporal validation, and performance evaluation

**DOI:** 10.64898/2026.08.03.26359583

**Authors:** Felipe Mendes Delpino, Daniela Magalhães, Igor Tona Peres, Tomoe Gusberti, Carlos José de Lima, Fernando A. Bozza, Otavio Ranzani, Leonardo S. L. Bastos

## Abstract

**Background:** Dengue continues to place a heavy clinical and organizational burden on health care systems, particularly during epidemics, when the high volume of cases adds pressure on triage, decisions regarding hospitalization, and the monitoring of patients at higher risk of severe outcomes. Despite the growing body of literature on dengue prediction, many studies still exhibit heterogeneity in outcomes, insufficiently detailed analytical designs, and a lack of validation.

**Objective:** To describe the protocol for a study on the development and validation of predictive models for the outcomes of hospitalization among reported cases and mortality among hospitalized patients.

**Methods:** A retrospective study will be conducted using secondary data from the Brazilian Notifiable Diseases Information System (SINAN), covering the period from 2017 to 2025. We will build two independent models: one to predict hospitalization among reported dengue cases and another to predict mortality among patients hospitalized for dengue. The protocol will follow TRIPOD+AI guidelines and include prior definition of eligible predictors, restriction to variables available at the clinically appropriate time of decision-making, handling of missing data, comparison between regression and machine learning algorithms, internal and temporal validation, and assessment of discrimination, calibration, and clinical utility.

**Conclusion:** The study aims to establish a transparent and reproducible analytical protocol to support the development of risk models that could be applied to clinical screening and surveillance for dengue.

## INTRODUCTION

Dengue is an arbovirus infection with a wide clinical spectrum, ranging from a self-limiting febrile syndrome to cases involving plasma leakage, severe bleeding, organ dysfunction, and death [1]. In endemic countries, its impact extends beyond individual morbidity to the organization of healthcare services, as large-scale epidemics increase the demand for triage, observation, hospitalization, and intensive care [1]. A revision of the clinical classification of dengue was proposed precisely to align the definition of severity with surveillance and actual patient management [2]. In recent Brazilian national studies, hospitalization and death remained significant outcomes on a large scale, reinforcing the need for clinically applicable risk stratification tools [3].

Although the literature on dengue prediction has grown in recent years, its primary focus has remained on clinical severity, rather than specifically on hospitalization or in-hospital mortality [4]. A recent systematic review showed a predominance of models for severity, a smaller number of models for mortality, and a scarcity of models directly focused on hospitalization [4]. The same review showed that calibration measures were reported infrequently and that most published models were rated at high risk of bias, mainly in the analysis domain: the handling of missing data was frequently inadequate or not described, the number of outcome events was small relative to the number of candidate predictors, and internal or external validation was often absent [4].

In clinical terms, hospitalization and mortality should not be treated as interchangeable outcomes [1,2]. The decision to admit a patient occurs at an earlier stage of the care pathway and depends on the information available during the initial assessment [1,2]. Mortality among hospitalized patients, on the other hand, corresponds to a later stage, in which the patient has already passed the threshold of greatest clinical severity and requires more intensive monitoring [1,3]. Separating these outcomes improves the clinical consistency of the study and reduces conceptual heterogeneity in the modeling [1,2].

How a prediction model is developed and reported matters as much as the outcome it targets, and studies of this kind are expected to follow established methodological and reporting standards [5,6]. TRIPOD+AI has become the primary guideline for reporting studies of predictive models based on regression and machine learning [5], TRIPOD-Cluster extends these recommendations to data organized by centers, regions, or time periods [7], and PROBAST, together with its update PROBAST+AI, provides structured tools for appraising risk of bias, methodological quality, and applicability [8,9]. Beyond risk of bias, published dengue models also leave a gap in what is reported about their results. Performance is still described mostly in terms of discrimination, even though highly discriminative algorithms can produce poorly calibrated absolute risks and therefore mislead clinical decisions [10,11]. Validation in independent samples or later periods remains uncommon, so the stability of performance across successive epidemic cycles is largely untested, and temporal leakage, inappropriate handling of missing data, and overfitting can artificially inflate apparent performance [6,8,9,12]. The assumed superiority of complex machine learning methods is also rarely supported, since methodological reviews have not demonstrated a consistent benefit of these methods over logistic regression in binary clinical models [13].

Here we describe a study protocol to develop and validate predictive models using Brazilian notifiable diseases surveillance (SINAN) data for two separate outcomes: hospitalization among reported dengue cases and mortality among hospitalized patients. The study will also characterize the eligible population and the temporal distribution of the outcomes; define and operationalize predictors available at the clinically appropriate decision-making time; compare regression and machine learning algorithms; assess discrimination, calibration, and clinical utility; and examine the temporal stability of performance over the study period.

## METHODS

This protocol describes a retrospective study for the development and temporal validation of prognostic models using routinely collected secondary data from the Brazilian Notifiable Diseases Information System (SINAN), covering eligible dengue records between 2017 and 2025. SINAN is the national compulsory notification system maintained by the Brazilian Ministry of Health, in which cases of dengue and other notifiable conditions identified in public and private health services throughout the country are registered at the individual level, together with demographic characteristics, dates of symptom onset and notification, signs and symptoms, laboratory results, hospitalization, and final case classification. It is therefore an epidemiological surveillance database covering the whole country, and not a clinical or laboratory record system restricted to hospitals. The study was designed in accordance with TRIPOD+AI [5]. If the final modelling strategy adopts a structure grouped by municipality, state, region, or epidemic period, the principles of TRIPOD-Cluster will also be incorporated [7]. Risk of bias and applicability will be appraised using PROBAST+AI [5,6,12].

The analytical workflow is organized around the six-step Reusable Analytical Pipelines for Infectious Diseases (RAPID) framework proposed by ISARIC, covering data cleaning, data preprocessing, modelling, model evaluation, validation, and visualization. RAPID is under active development, and this protocol refers to its current publicly available version. Structuring the protocol in this way situates the study within ISARIC’s analytical ecosystem, favors reproducibility, and allows individual components of the pipeline to be reused or replaced without compromising the overall design. Figure 1 summarizes the sequence of steps from data extraction to model output.

**Figure 1.**
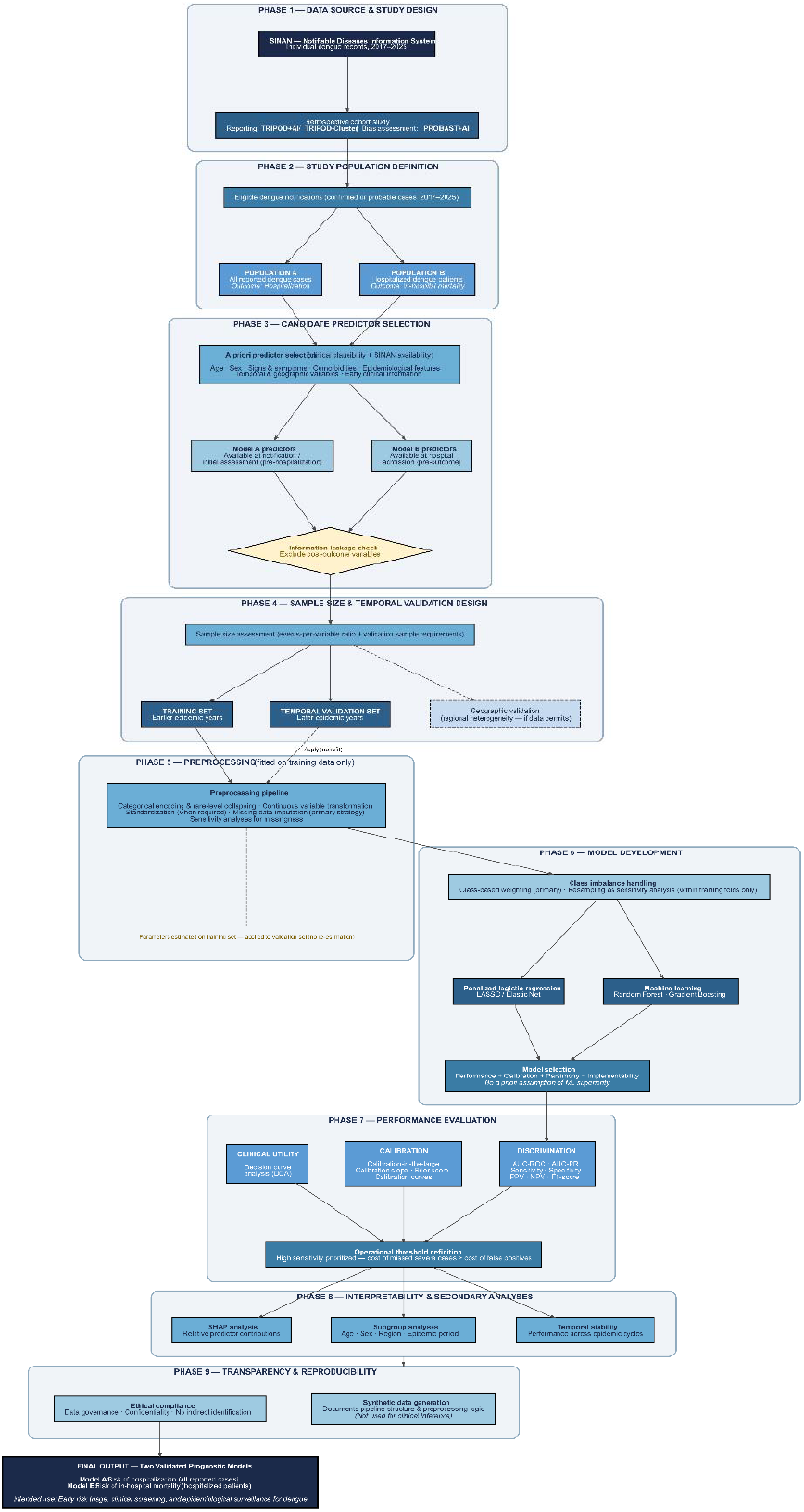
Overview of the analytical pipeline for the development and temporal validation of predictive models for hospitalization and in-hospital mortality in dengue using SINAN data (2017–2025)

Two analytical populations will be defined and kept independent throughout the pipeline: all reported dengue cases for the hospitalization model, and patients hospitalized for dengue for the mortality model. This separation reflects two distinct clinical questions and prevents extrapolation of a single model across different points of the care pathway [1–4]. The outcome of the first model is hospitalization, under the assumption that the time of prediction must precede admission. The outcome of the second model is death among hospitalized patients. Rather than adopting fixed 28-day or 60-day horizons, which are long in relation to the natural history of dengue, the primary outcome will be death occurring during the hospital episode; the interval between admission and death will be described, hospital stays of up to 15 days will be used as the reference window for in-hospital mortality, and death from any cause recorded at case closure, without a time window, will be examined as a secondary outcome.

Eligibility will follow the case definitions used in routine surveillance. All notified dengue cases closed as confirmed, by laboratory or by clinical-epidemiological criteria, will be eligible; discarded cases and records whose final classification remains inconclusive will be excluded. No age restriction will be applied, so that children, adults, and older adults are represented as they appear in routine care; age will be modelled as a candidate predictor and used in pre-specified subgroup analyses. Pregnancy, reported comorbidities, and documented co-infection with other arboviruses will not be exclusion criteria and will be retained as candidate predictors whenever recorded. Records will be excluded when the outcome of interest is missing, when the date of symptom onset or of notification is missing, or when the sequence of dates is incompatible with the modelled decision point. For the mortality model, eligibility will be restricted to records with a documented hospitalization, and deaths attributed at case closure to causes unrelated to dengue will be examined in sensitivity analyses.

Candidate predictors will be defined a priori based on clinical plausibility, availability in SINAN, and potential operational utility, including age, sex, signs and symptoms, recorded comorbidities, epidemiological characteristics, and temporal and geographic variables. The final selection will adhere to the principle of clinical temporality: the hospitalization model will use only information available up to notification or initial assessment, and the mortality model will use information available up to admission. Variables recorded after the outcome, or clearly resulting from it, will be excluded to prevent information leakage [5,6,8,9,12].

Data cleaning will precede any modelling decision, in line with the first step of the RAPID framework, which includes the treatment of duplicate notifications known to occur in SINAN. When the same patient is reported more than once, duplicate records will be identified through key-based deduplication, and we will retain the earliest or the most complete entry according to the clinical question. Variables with incompatible units or coding across years will be harmonized, and features with zero or near-zero variance will be removed using a frequency-ratio criterion, since they add no predictive information and may destabilize some algorithms. The extent and pattern of missing data will be quantified for every candidate variable, and the likely missingness mechanism will be assessed before imputation, since treating data that are not missing at random as if they were can introduce bias that no downstream step corrects.

The preprocessing step will be fitted on the training data and applied to validation data without re-estimation. Categorical predictors will be recoded with aggregation of rare categories where clinically justified and transformed through one-hot encoding; continuous predictors will be inspected for skewness and, when needed, transformed and standardized. Collinearity will be examined using pairwise Pearson correlations and variance inflation factors for continuous predictors, and Cramér’s V for associations between categorical predictors. Temporal variables relevant to dengue will be encoded with attention to their cyclical nature: month and epidemiological week will be represented through sine and cosine transformations, preserving the seasonality central to arbovirus dynamics in Brazil. Predictors will additionally be organized into conceptual blocks, namely demographic, clinical, temporal or seasonal, and geographic variables, so that the incremental predictive contribution of each block can be assessed alongside that of individual variables; this is particularly relevant for season and for region or municipality, whose contribution is otherwise difficult to separate from patient-level information. Feature selection will combine filter approaches with embedded methods such as LASSO and Elastic Net. Wrapper strategies, including recursive feature elimination guided by the algorithm to be fitted, may be used for models that do not perform selection internally, whereas ensemble methods that already incorporate feature selection as part of model fitting will be used without an additional selection step. Missing data will be handled primarily through Multiple Imputation by Chained Equations (MICE), with median or mode imputation retained as a sensitivity analysis.

The sample size will be determined based on the number of available events and the number of candidate parameters, recognizing that studies developing binary models must ensure an adequate number of participants and events to reduce overfitting and instability in the estimates [14]. Since the accuracy of the evaluation in independent samples also depends on the number of observed events, the validation step will be interpreted in accordance with the recommendations for sample size in external validation of binary models.

Both outcomes are binary, and for each of them regression models will be compared with supervised machine learning algorithms suited to binary classification, including regularized regression, tree-based ensembles such as random forests, and gradient boosting methods, provided they are compatible with the final database structure and with the events-to-complexity ratio. These families were chosen because together they span the range from a transparent and easily deployable benchmark to flexible algorithms capable of capturing non-linearities and interactions among signs, symptoms, and temporal or geographic variables, while remaining computationally feasible on tabular surveillance data of this size. Specific implementations, hyperparameter grids, and tuning procedures will be reported in full in the supplementary material of the final paper: the strategy is data-driven, and the final specification depends on characteristics of the assembled dataset that can only be observed once the analysis is carried out. A stacking ensemble, combining the predictions of the best-performing models through a second-level learner, will additionally be evaluated as an exploratory analysis [15]. The choice of the final model will not assume the automatic superiority of more complex approaches. Methodological reviews have shown that, in many clinical contexts, machine learning models did not outperform logistic regression when properly compared [13]. For this reason, performance, calibration, parsimony, and interpretability will be prioritized over marginal gain in discrimination [10,11,13].

Since both outcomes may be unbalanced, particularly mortality, class imbalance will be addressed primarily through class weights inversely proportional to outcome frequency and other approaches compatible with cost-sensitive learning, which preserve the observed outcome prevalence and therefore the interpretation of predicted probabilities as absolute risks. Resampling techniques, namely random undersampling of the majority class and synthetic minority oversampling, will be treated as sensitivity analyses and applied only within the training folds, never to validation or test data, to reduce artificial inflation of performance and preserve a more realistic estimate of out-of-sample performance [5,6,12]. Whenever resampling or class weighting is applied, predicted probabilities will be recalibrated before performance is reported, so that discrimination is not improved at the expense of calibration.

Model evaluation will rely on repeated k-fold cross-validation within the training window combined with a held-out temporal test set [16,17]. The primary validation will be temporal, with training on earlier years and testing on subsequent periods within the 2017–2025 window, simulating prospective use and exposing temporal drift in performance [12]. If the data structure permits, validation by geographic units will also be explored, with emphasis on regional heterogeneity, which is particularly relevant in distributed surveillance databases in Brazil [7,12]. Uncertainty around performance metrics will be quantified through non-parametric bootstrapping [16], and sensitivity analyses will test robustness to imputation strategy, operational thresholds, and alternative outcome definitions.

Performance will be evaluated based on discrimination, calibration, and clinical utility. Discrimination will include the Area Under the Receiver Operating Characteristic Curve (AUROC), the Area Under the Precision-Recall Curve (AUC-PR), and classification threshold-dependent metrics such as sensitivity, specificity, positive predictive value, negative predictive value, and F1-score [10]. Calibration will include calibration-in-the-large, calibration slope, Brier score, and calibration belts with confidence intervals [10,11]. Clinical utility will be examined using decision curve analysis [17]. The choice of these metrics reflects the understanding that the utility of a model depends on adequate separation between cases and non-cases, accuracy of predicted probabilities, and clinical relevance of decision thresholds [10,11,17].

The operational thresholds will be defined transparently and compatibly with the intended use of the models as tools for early risk triage and stratification. For both outcomes, high sensitivity will be prioritized, with reporting of the corresponding trade-off in specificity and false positives, since in dengue scenarios the clinical cost of missing cases with poor evolution tends to outweigh a moderate increase in false-positive classifications [1,3,17].

The interpretability of tree-based and boosting models will be explored using SHAP analyses [18,19]. The protocol also provides for secondary analyses by age, sex, geographic region, and epidemic period, given that the literature shows that factors such as age, comorbidities, warning signs, and laboratory variables are frequently associated with unfavorable clinical progression in dengue [20–22]. All visual outputs will be generated exclusively from aggregated data frames, in line with the RAPID principle that patient-level information should not leave the pipeline. When applicable, these outputs will include forest plots for regression effect sizes, calibration curves, correlation and confusion heatmaps, upset plots for co-occurring comorbidities and warning signs, and Sankey diagrams for the flow of patients from notification through hospitalization to outcome, making the two-model architecture visually explicit.

As part of the strategy for transparency and reproducibility, the pipeline will be implemented as open-source code in a public repository with version control, code review, unit and integration tests, and documentation at the function and tutorial level, following RAPID coding standards. Because SINAN microdata are publicly available, the repository will also include the extraction and preparation scripts required to rebuild the analytical dataset from the original public files, so that the whole analysis can be reproduced without access to restricted data. Because the study uses secondary surveillance data, it will comply with the ethical and regulatory requirements applicable to the governance and use of administrative health databases. The presentation of the results will aim to preserve confidentiality, prevent indirect identification, and prioritize transparent communication of analytical limitations.

## DISCUSSION

This protocol was structured around two distinct outcomes, since hospitalization and in-hospital mortality represent different clinical and epidemiological decisions. The literature on dengue often conflates severity, admission, need for intensive care, and death into a single analytical continuum [4,20–23]. Although this may be understandable from a pathophysiological perspective, this grouping reduces the clinical utility of the model when the intention is to support real-world decisions at specific points in care [2,5,6].

The decision to separate outcomes improves the consistency of the modelling. A useful model for hospital admission should operate using information available prior to admission [1,2]. In contrast, a model for in-hospital mortality needs to reflect a more severe clinical population, in which the objective is not to decide on admission but to identify deterioration and extreme risk among patients already hospitalized [3,22]. This distinction also facilitates a clearer interpretation of predictive variables and operational thresholds [5,6].

The choice of temporal validation is another key aspect of this protocol. In dengue, epidemic intensity, circulating serotypes, age profile, service organization, and regional disease patterns may vary over time [1,3]. The 2017–2025 window therefore offers methodological potential for examining the temporal stability of performance [3,12].

A significant portion of the clinical literature continues to focus solely on AUROC or other classification metrics [4,10,11]. Highly discriminative models can nonetheless produce poorly calibrated absolute risks and, as a consequence, lead to inappropriate decisions [11]. In a model designed to support hospitalization decisions or monitor mortality risk, this matters because small distortions in individual probability can shift clinical thresholds for referral and escalation of care [11,17].

Organizing the protocol within the RAPID framework contributes beyond this particular study. By documenting the pipeline against the six RAPID steps and sharing the code openly, this study can serve as a reference implementation that other groups may adapt to different infectious diseases, data sources, and outbreak contexts.

Predictive studies on dengue have shown that relatively simple scores and models can achieve useful performance in specific scenarios [21,23], and machine learning models have also shown potential in this field [20]. Nevertheless, the methodological literature indicates that the superiority of machine learning over logistic regression should not be assumed as a rule, especially when comparison, validation, and reporting are not conducted rigorously [13]. For this reason, the choice of the final model in this study will depend on validated performance, calibration, temporal stability, and feasibility of implementation, and not merely on the best apparent metric during training [10,11,13].

In summary, this protocol proposes a methodological strategy for a clinically relevant problem. By clearly separating outcomes, specifying the predictive time point, prioritizing temporal validation, incorporating robust calibration assessment, and documenting the pipeline as a reusable RAPID, the study aims to produce more defensible and potentially more useful models for large-scale dengue screening, monitoring, and clinical surveillance.

## Data Availability

All data produced in the present work are contained in the manuscript.

